# Behavior-change interventions to improve antimicrobial stewardship in human health, animal health, and livestock agriculture: A systematic review

**DOI:** 10.1101/2023.01.04.23284191

**Authors:** Jessica Craig, Rachel Sadoff, Sarah Bennett, Felix Bahati, Wendy Beauvais

## Abstract

Antimicrobial resistance (AMR) is an economic, food security, and global health threat that is driven by a multitude of factors including the overuse and misuse of antimicrobials in the human health, animal health, and agriculture sectors. Given the rapid emergence and spread of AMR and the relative lack of development of new antimicrobials or alternative therapies, there is a need to develop and implement non-pharmaceutical AMR mitigation policies and interventions that improve antimicrobial stewardship (AMS) practices across all sectors where antimicrobials are used. We conducted a systematic literature review per the Preferred Reporting Items for Systematic Reviews and Meta-Analyses guidelines to identify peer-reviewed studies that described behavior-change interventions that aimed to improve AMS and/or reduce inappropriate antimicrobial use in the human health, animal health, and livestock agriculture stakeholders. We identified 301 total publications – 11 in the animal health sector and 290 in the human health sector – and assessed these interventions using metrics across five thematic areas- (1) antimicrobial use (AMU), (2) adherence to clinical guidelines, (3) AMS, (4) AMR, and (5) clinical outcomes. The lack of studies in the animal health sector precluded a meta-analysis. Among studies in the human health sector, 35.7% reported significant (p<0.05) pre- to post-intervention decreases in AMU, 73.7% reported significant improvements in adherence of antimicrobial therapies to clinical guidelines, 45% demonstrated significant improvements in AMS practices, 45.5% reported significant decreases in the proportion of isolates that were resistant to antibiotics or the proportion of patients with drug-resistant infections across 17 antimicrobial-organism combinations, and few studies reported statistically significant changes in clinical outcomes. We did not identify any overarching intervention type nor characteristics associated with successful improvement in AMS, AMR, AMU, adherence, nor clinical outcomes.

## Introduction

Antimicrobial resistance (AMR) is an economic, food security, and global health threat (1–3). Globally, there were an estimated 4.95 million and 1.27 million human deaths associated with or directly attributable to drug-resistant bacterial infections, respectively, in 2019 (4). The Centers for Disease Control and Prevention estimates that the annual cost to treat hospital- and community-acquired drug-resistant bacterial infections in the US alone is $4.6 billion due to prolonged hospital stays, more complex healthcare needs, and treatment with second- and third-line antimicrobial therapies (5). In addition to the human health burden, AMR among pathogens affecting animals has adverse animal welfare impacts and imposes additional treatment costs on animal owners, ultimately leading to increased costs of food production. The emergence of new drug-resistant pathogens may also hinder global food and animal trade (3,6). Researchers estimate that AMR may cause a 7.5 percent decline in global livestock production by 2050 (7).

Drug resistance is driven by a multitude of factors including the overuse and misuse of antimicrobials in the human health, animal health, and agriculture sectors. Although antimicrobial use (AMU) and consumption (AMC) surveillance is limited in the human and animal health sectors, available evidence demonstrates that AMU is increasing globally. Analysis of AMC rates, approximated from data on national-level antimicrobial sales data, indicated a 39% increase in per capita AMC in the human health sector between 2000 and 2015, rising from 11.3 to 15.7 daily defined doses per 1,000 people per day (DIDs) (8). Concerningly, the use of World Health Organization-designated “Watch” antibiotics has increased faster than “Access” antibiotics. Between 2000 and 2015, there was a 90.9% increase in the global per capita consumption of “Watch” antibiotics, from 3.3 to 6.3, compared to a 26.2% increase in the global per capita consumption “Access” antibiotics which rose from 8.4 to 10.6 DIDs in the same period (9). Previous studies show clinically inappropriate AMU rates as high as 55% in South Africa, 88% in Pakistan, and 61% in China in the human health sector; however, emerging evidence suggests there is significant heterogeneity in AMU practices across countries and clinical settings (10–13). Best available evidence suggests that AMU in the animal health sector is rising as the global demand for animal products increases (14). In 2017, there were an estimated 93,309 tons (95% confidence interval [CI]: 64,443-149,886) of antimicrobials sold for use in chicken, cattle, and pig systems across 41 countries. In the same year, AMC in the aquaculture sector was estimated at 10,259 tons across six fish species in 33 countries (15). By 2030, AMU is projected to rise to 104,079 tons among chicken, cattle, and pigs and to 13,600 tons among fish (14,15).

Concerningly, the emergence and evolution of drug-resistant microorganisms is outpacing the development of new antimicrobial agents. In the past 50 years, no new antimicrobial agents active against gram-negative bacteria have been brought to market, and only 5 of the 20 major global pharmaceutical companies are engaged in antimicrobial research and development (R&D) (16). Investment in antimicrobial R&D is not considered lucrative given the challenges in antimicrobial discovery, rapid introduction of generic formulations, and the speed at which microorganisms develop resistance to antimicrobials. Moreover, new antimicrobials are likely to be reserved for special or emergency use to preserve their efficacy; therefore, there would paradoxically be less demand for these drugs.

While AMR poses an increasing public health threat, the number of deaths that could have been prevented by improving access to antimicrobials, about 5.7 million in low- and middle-income countries (LMICs), exceeds the morbidity and mortality burden from AMR indicating that access to clinically appropriate antimicrobials for the treatment of infectious diseases remains a critical issue (16).

Given the health and economic impacts of AMR, challenges around access to appropriate antimicrobial therapies, and the lack of antimicrobial R&D, there is an urgent need to develop and implement impactful non-pharmaceutical AMR mitigation policies and interventions that improve antimicrobial stewardship (AMS) practices in the human health, animal health, and agriculture sectors. There is a growing body of literature describing such non-pharmaceutical AMS interventions which include education, training, and health information campaigns to improve awareness and knowledge among technical stakeholders and the public; installing AMS committees in healthcare facilities to approve and/or review prescription practices; and developing and implementing standard clinical treatment guidelines or clinical treatment algorithms which provide clinicians with an evidence-based resource to guide clinical decision making around AMU and prescription (17–19). Despite this emerging evidence, there remains no consensus on what constitutes the most impactful or cost-effective AMS practices across various sectors and settings. Therefore, the purpose of this study was to conduct a systematic literature review and meta-analysis to identify behavior-change interventions aimed at improving AMS and AMU across the human health, animal health, and agriculture sectors to identify gaps in the evidence base or to describe trends towards best AMS practices in various resource and income settings.

## Materials and Methods

This systematic review and meta-analysis followed the Preferred Reporting Items for Systematic Reviews and Meta-Analyses (PRISMA) guidelines (20). We searched PubMed, Web of Science, Embase, Centre for Agriculture and Bioscience International (CABI), and the Cochrane Database of Systematic Reviews for peer-reviewed studies that described behavior-change interventions that aimed to improve AMS and/or reduce inappropriate AMU or AMC among human health, animal health, and livestock agriculture stakeholders including but not limited to health providers, patients, farmers, or animal owners (21–25). Two searches were conducted; the first on 15 June 2021 identified studies published prior to 15 June 2021 and a second search conducted on 31 August 2022 identified studies published between 15 June 2021 and 31 August 2022 (inclusive). The following search string was used to search the title and abstracts of published entries within each database: (intervention OR trial) AND (antimicrobial” OR antibiotic) AND (use OR stewardship OR consumption OR prescription).

Two researchers independently screened returned entries for eligibility in a two-step process: a review of the title and abstract followed by full-text review, according to the following criteria: (1) Only scientific articles published in peer-reviewed journals were included; pre-prints, course completion papers or theses, and internal reports or policy briefs were excluded. (2) Papers published in any language and written on interventions conducted in any country were included. (3) All study types including randomized controlled trials and observational studies (including pre/post study designs) were included. Review articles were screened for additional relevant studies. (4) Studies were included if they reported on at least one outcome which fell into three general categories: (a) AMU or AMC; (b) adherence/compliance of antimicrobial therapy to facility, national, or international clinical treatment guidelines; and (c) rates of specific stewardship practices such as the intravenous to oral therapy conversion rates, frequency of obtaining specimen cultures for pathogen identification and/or antimicrobial susceptibility tests. For studies that reported on at least one primary outcome, we also extracted data for this study’s secondary outcomes which included the incidence or prevalence of drug-resistant infections; length of stay in the healthcare facility or specific unit (such as intensive care); health outcomes such as patient/animal mortality or rehospitalization rates; and the economic or cost impact of interventions. (5) Non-relevant papers that did not describe interventions aimed at changing behaviors to optimize AMU, AMC, or AMS in the human health, animal health, or livestock agriculture sectors or those that did not report on our primary outcomes of interest were excluded. Therefore, studies such as those that reported solely on changes in knowledge, attitudes, and perceptions regarding AMR or AMU were excluded as were studies that assessed drug or drug regimen efficacy. (6) Studies were also excluded if the full text was unavailable or unobtainable, if results and statistics were not presented numerically (such as only presented graphically), or if insufficient data was reported to conduct statistical analyses such as to calculate odds ratios or risk ratios or conduct t-tests or Chi-square tests. At each stage, two reviewers independently screened entries for eligibility, identified discrepancies, and re-evaluated articles until a consensus was reached. Duplicates were identified when results from each search were combined into a single database.

Data extraction was conducted by one researcher (JC, SB, FB) and reviewed by a second (JC, SB, and FB); data was compiled using Microsoft Excel software. We used a standardized spreadsheet (S4 file) to extract data from each publication that met our inclusion criteria, including the publication year, country and setting on the intervention, type of evaluation, type of study, description of intervention, intervention start and end date and duration, total sample size and unit of sample size, the study’s outcome, and all relevant numerical data reported. The income classification of the country where the intervention occurred, per the World Bank’s 2021 classification, was also extracted (26). We classified studies as being either single or multi-center and we classified interventions either single or bundled (meaning more than one type of intervention occurred), based on the publication. Duration of intervention was computed, if not provided in the study, using the intervention start and end dates.

For the meta-analysis, we created syntheses based on three criteria: type of statistic reported, study type, and outcome variables. First, studies were categorized by type of statistic (rates or proportions) reported. Then studies were categorized by study type or data analysis/reporting methodology. These categories included: pre/post observational studies; randomized or non-randomized control trials (RCTs) that compared a control arm to an experimental/intervention arm; randomized or non-randomized control trials that compared one intervention arm to another intervention arm; and time series studies. We then categorized the studies by outcome. For all studies, odds ratio, rate ratio, 95% confidence interval, chi-square statistic, Wilcoxon signed-rank test, and independent or dependent t-test statistics were calculated, as relevant, to compare the study arms or pre- to post-intervention changes according to the study outcomes. Bivariate analyses and linear and logistic regression analyses were conducted across studies to identify trends in intervention success in various settings. For logistic regression analyses for the pre- to post-intervention studies, a categorical variable was created with 3 categories indicating if there was a significant increase, significant decrease, or no statistically significant change in pre- to post-intervention outcomes. Backwards stepwise elimination process based on likelihood ratio tests was used to build final regression models. The Quality Assessment Tool for Quantitative Studies was used to assess study quality and risk of bias (27). For the quality assessment, each study was independently reviewed by two researchers with a third to resolve disputes and compute a consensus quality score.

## Results

Our review returned a total of 154,106 publications of which 4,893 were duplicates; 148,639 were determined to be irrelevant based on the title and abstract review; and 36 were identified as review articles. Following full-text review, a total of 301 publications – 11 in the animal health sector and 290 in the human health sector (S1 and S2 files) – met the study’s inclusion criteria and underwent data extraction (Fig 1). Per the Quality Assessment Tool for Quantitative Studies, 4 of 11 studies that described interventions in the animal health or agriculture sectors (36.4%) were determined to have strong overall quality ratings, 3 (27.3%) had moderate quality ratings, and 4 (36.4%) had weak quality ratings while 129 of the 290 studies (44.5%) that described interventions in the human health sector were determined to have strong overall quality, 115 (39.7%) had moderate overall quality, and 46 (15.9%) had low overall quality scores (S3 file). A common reason for low quality scores was a high risk of selection bias or failing to control for potential cofounders.

**Fig 1.**
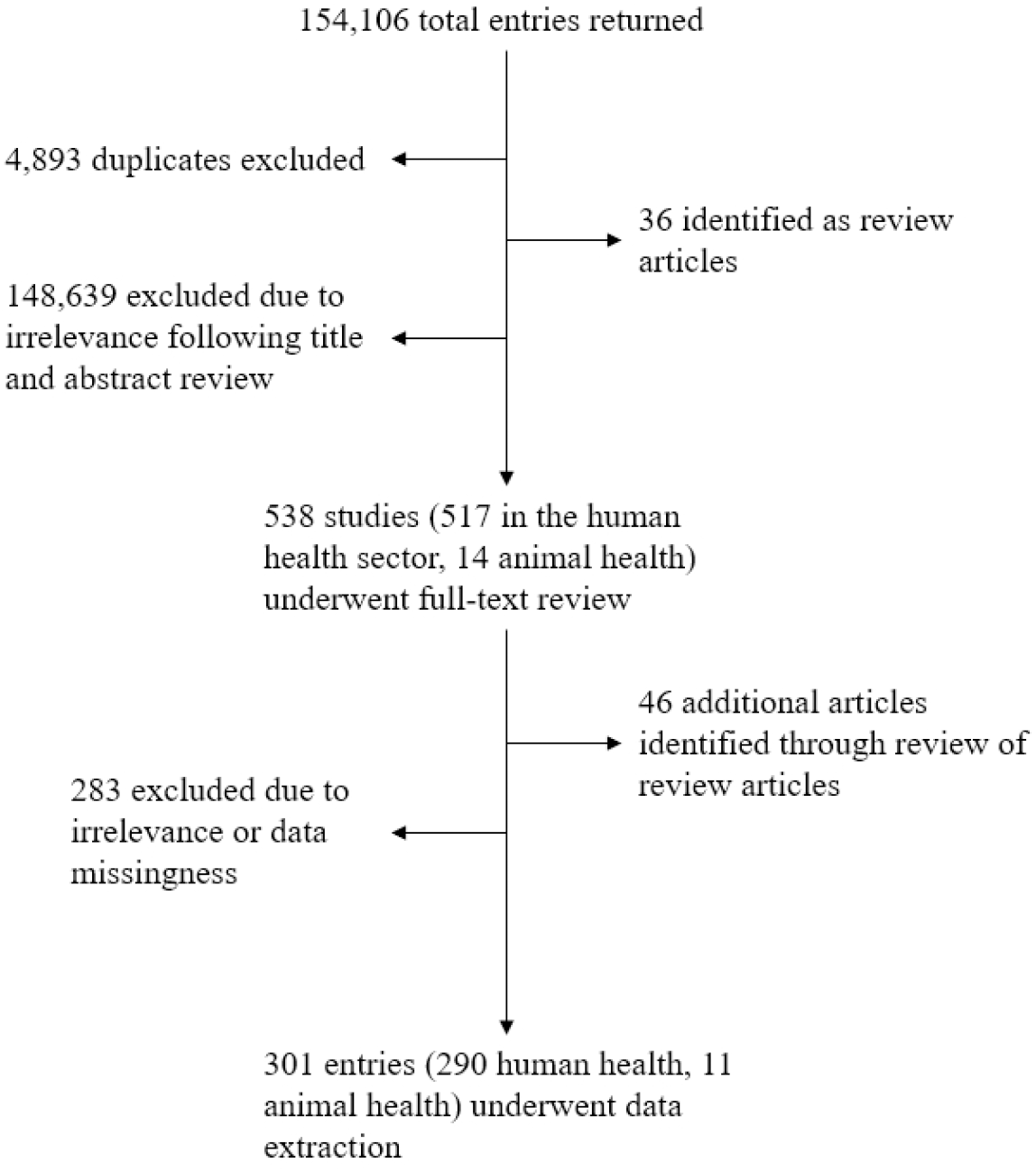
Flow diagram of systematic literature review describing behavior change interventions to improve antimicrobial use and stewardship in the animal and human health sectors.

### Animal health and agriculture sectors

Only 11 studies conducted in the animal health or agriculture sector met our study’s inclusion criteria, and given heterogeneity across the study types, outcomes, and types of statistics reported, a meta-analysis was not possible. Seven of the studies (63.6%) assessed the impact of interventions among food-producing animals including swine (n=3), poultry (n=1), dairy cows (n=2), and calves (n=1) (28–34). Four studies (36.4%) assessed interventions in companion animals (35–38). Year of study publication ranged from 2014 to 2021. Across the 11 studies, 8 countries were represented (Germany, Sweden, Netherlands, Belgium, France, Switzerland, the United Kingdom, and Vietnam). Interventions ranged from 5 months in duration to 5 years. Five studies (45.5%) implemented bundled interventions while the remaining 7 studies (63.6%) tested single interventions. Four studies (36.4%) implemented online AMS tools that either provided guidance on AMU and AMS or restricted antimicrobial prescriptions. Other interventions included veterinary guidance on husbandry, biosecurity, or AMU; educational programs; review and feedback of AMU; and programs to implement infection prevention and control measures. Overall AMU decreased significantly (p value < 0.05) for all but one study which implemented an online stewardship tool among calf farmers in Switzerland (29). Three studies assessed pre- to post-intervention changes in animal health status and productivity and documented either no change or an improvement in mortality, animal weight gain, or feed conversion ratio associated with the intervention (28,32,33). One study that implemented a voluntary AMS program among pig farmers in France documented a 90% reduction in cephalosporin usage between 2010 and 2016 and demonstrated a reduction in drug-resistance among commensal and pathogenic *Escherichia coli* isolates sampled during the same period (31).

### Human health sector

Of the 290 studies that assessed behavior-change interventions in the human health sector, 40 (13.7%) were RCTs, 237 (81.7%) were pre/post observational studies, and the remaining 13 (4.5%) followed alternative study designs such as historical cohort study. Ninety-seven studies (33.4%) were retrospective, 129 (44.5%) were prospective, 49 studies (16.9%) utilized both retrospective and prospective evaluations, and the remaining studies did not specify the evaluation type utilized to assess the intervention. Year of publication ranged from 2001 (n=2) to 2021 (n=44). Studies were most commonly conducted in high-income countries (n=228 studies, 78.6%) with 32, 21, and 3 studies (11.0, 7.2, 1.0%, respectively) conducted in upper-middle, lower-middle, and low-income countries, respectively. A total of 49 countries were represented across the studies that met our inclusion criteria; USA (n=110 studies), Canada (n=16), Japan (n=15), Spain (n=15), China (n=12), and Italy (n=10) were the most represented. Academic/teaching hospitals or tertiary care facilities were the most common setting for intervention implementation.

Given the smaller number of studies that utilized RCTs designs and the heterogeneity across data and outcomes, a meta-analysis of RCTs was not possible; therefore, we focus our meta-analysis on interventions evaluated through pre/post observational study designs.

#### Theme 1: Antimicrobial use

Of 290 studies that described interventions in the human healthcare setting and met our eligibility criteria, we extracted AMU data, quantified as a proportion of *patients* who received an antimicrobial before and after the study intervention, from 42 studies (Fig 2). The denominator (i.e. *patients*) was defined differently in each study e.g. patients presenting at a primary healthcare facility, patients in a long-term care facility. Amongst the 15 of 42 studies (35.7%) that found a statistically significant (p value <0.05) pre- to post-intervention decrease in AMU; the average decrease in the proportion of patients receiving various antimicrobial agents or classes was 10.4% (Standard deviation [SD]: 16.6) and the range was 3.1 to 58%. Four of these studies described interventions in lower-middle-income countries (Bangladesh, India, n=2, Iran), two in upper-middle-income countries (China, Malaysia), and 9 in high-income countries (Finland, Hong Kong, Netherlands, Spain, UK, USA, n=4). The clinical settings involved in these interventions included a veteran’s hospital, a university dentistry practice, a long-term care facility, a pediatric hospital, primary healthcare or community clinics (n=4 studies), secondary or tertiary hospitals (n=5 studies). Across these 15 studies, 7 (46.7%) assessed single interventions and the remaining 8 (53.3%) tested bundled interventions; intervention duration ranged from 6 weeks to 3 years. Audit or review and feedback of provider prescribing or AMU practices (n=7 studies, 46.7%) and education (n=6, 40%) were the most common intervention types implemented. Four studies assessed the impact of introducing various online or application-based diagnostic and clinical support tools, three studies introduced prescribing restrictions or pre-authorization requirements, two studies involved the development and/or introduction of a clinical treatment guideline, and one study implemented a diagnostic stewardship strategy to withhold laboratory results if there were no white blood cells or bacteria identified on microscopy.

**Fig 2.**
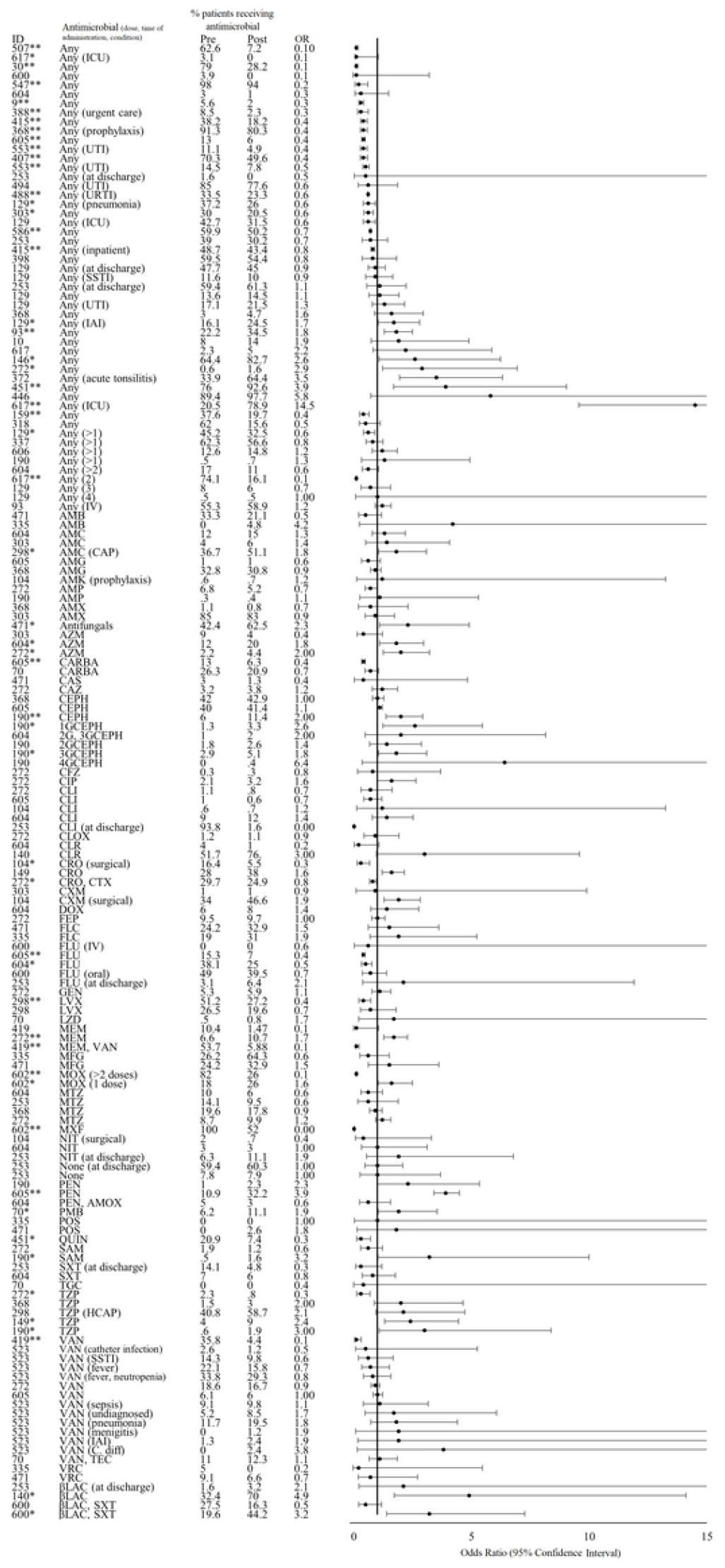
Effect of interventions on antimicrobial use (proportion of patients receiving antimicrobials). *Indicates significance <0.05. **Indicates significance <0.001. Antimicrobial abbreviations: AMK-amikacin, AMX-amoxicillin, AMC-amoxicillin-clavulanic acid, AMP-ampicillin, CARB-carbapenems, CTX-cefotaxime, CIP-ciprofloxacin, FOF-Fosfomycin, IPM-imipenem, LVX-levofloxacin, MDR-multi-drug resistant, MEM-meropenem, MET-methicillin, NIT-nitrofurantoin, TZP-piperacillin-tazobactam, SXT-trimethoprim-sulfamethoxazole, 3G-CEPH-third-generation cephalosporins. Other abbreviations: HCAP-healthcare associated pneumonia, IV-intravenous, CAP-community-acquired pneumonia, ICU-intensive care unit, IAI-intraabdominal infection, UTI-urinary tract infection, SSTI-skin or soft tissue infection.

Six of 42 of the studies (14.3%) that reported on the proportion of patients who received an antimicrobial documented a statistically significant (p value <0.05) increase in AMU after the intervention; the percentage increase averaged 10.01% (SD: 14.5) and ranged from 1.1 to 58.4%. All six of these studies described interventions in high-income countries (Denmark, Japan, Australia, USA, Spain, and Ireland); clinical settings of interventions included secondary or tertiary hospitals (n=3 studies), academic or university hospitals (n=2), and a children’s hospital. The majority of these 6 studies implemented bundled interventions (n=5, 83.3%); 4 included an audit or review and feedback activity; 3 involved the development and/or implementation of a clinical treatment guideline, protocol, or policy; 2 involved AMS education or training; 1 study introduced an AMS application; and another study introduced a rapid blood culture diagnostic test. Intervention duration ranged from 2 months to 45 months.

Seven studies (16.7%) reported simultaneous increases and decreases in different antimicrobial agents and classes that were statistically significant, ranging from a 21.3% pre- to post-intervention increase to a 56% decrease. For example, Leis *et al*., reported a significant reduction in the proportion of patients prescribed fluoroquinolone (15.3 to 7%, p<0.001) and carbapenems (13 to 6.3%, p<0.001) and an increase in the proportion of patients prescribed penicillin (10.9 to 32.2%, p<0.001) after an intervention that implemented a point-of-care β-lactam allergy skin test among other AMS activities (39). Another intervention described by Yogo *et al*. aimed to reduce broad-spectrum antibiotic prescribing rates and treatment durations at time of hospital discharge by introducing institutional guidance for oral step-down antibiotic selection and duration of therapy and implemented a pharmacy audit and real-time recommendations of discharge prescriptions (40). This study documented a significant pre- to post-intervention decrease in fluoroquinolone use (38.1 to 25%, p=0.001) and a significant increase in the proportion of patients prescribed azithromycin (12 to 20%, p=0.01). Across all datapoints reported in these 7 studies, there was an average 1.8% decrease in the proportion of patients receiving antimicrobials.

#### Theme 2: Appropriateness of therapy and adherence to treatment guidelines, protocols, and policies

We extracted data from 38 studies that described pre- to post-intervention changes in the adherence/concordance of AMU or prescribing practices with local, national, or international treatment guidelines or protocols (Fig 3). Of those, 28 studies (73.7%) reported a significant improvement in appropriateness and/or adherence while 1 study (2.6%) reported a significant decline. Most studies that reported improvements in appropriateness of therapy or adherence to guidelines were conducted in high-income countries (n=20, 71.4%, Australia; Canada, n=2; Denmark; Germany; Greece; Ireland; Israel; Italy, n=3); Netherlands; USA, n=8) with 2 studies in upper-middle-income (Malaysia, South Africa), 5 in lower-middle (Egypt; India, n=2; Kenya; Pakistan), and 1 in low-income countries (Nepal). Most studies assessed bundled interventions (n=19, 67.9%) in secondary, tertiary, or academic/teaching hospital settings. Audit or review and feedback; the development of implementation of clinical treatment guidelines, protocols, or policies; and education or training were the most common interventions implemented as the sole intervention or in combination with other intervention activities in 18 (64.3%), 16 (57.4%), and 14 (50%) studies, respectively. Intervention duration ranged from 3 months to 6 years.

**Fig 3.**
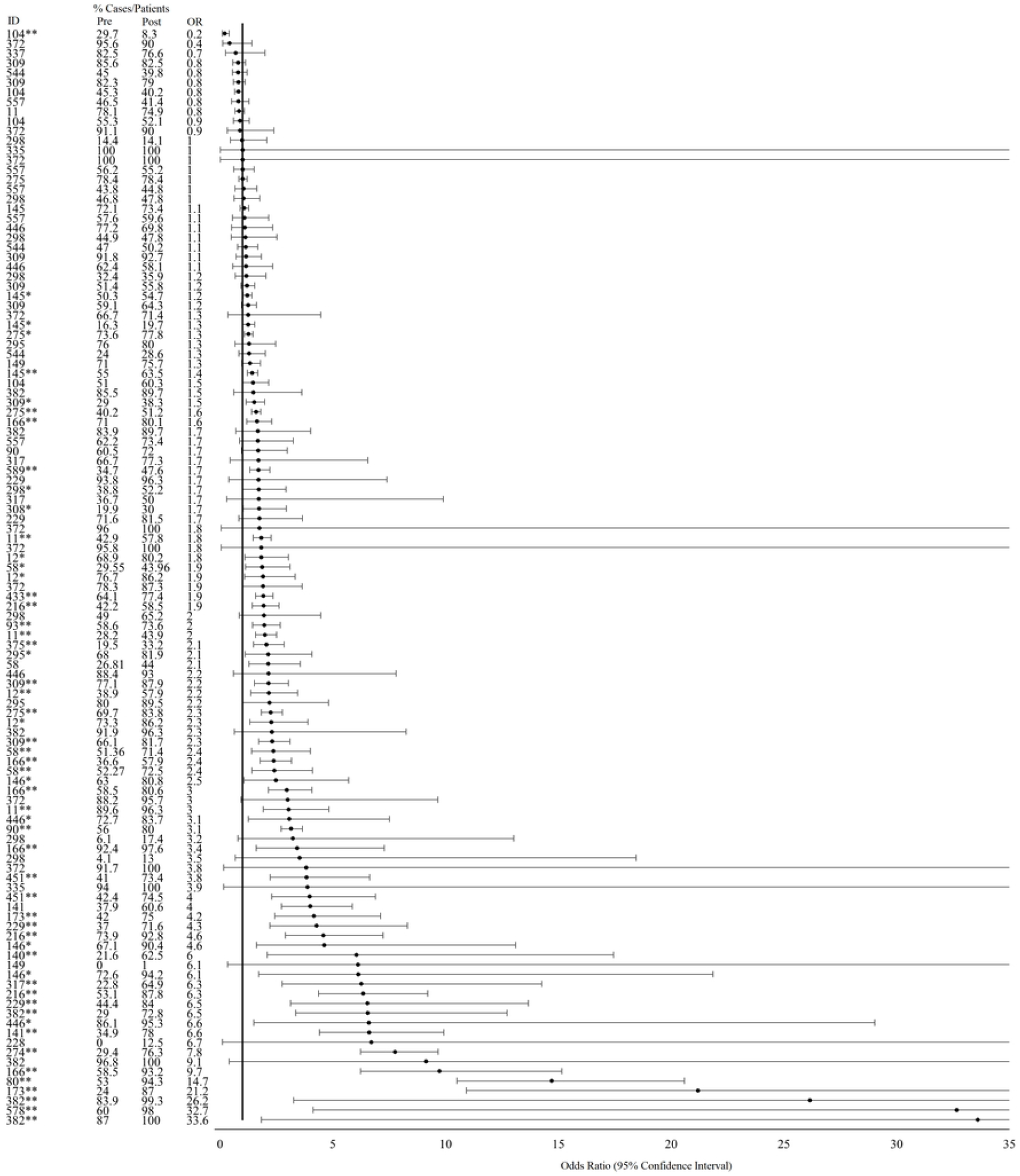
Effect of interventions on appropriateness of antimicrobial therapy or adherence to clinical guidelines (including drug selection, dosage, duration, and timing of administration). *Indicates significance <0.05. **Indicates significance <0.001.

The average increase in the proportion of cases or patients treated with appropriate or guideline-concordant antimicrobial therapies across all studies and indicators where a statistically significant change was reported was 20.8% (SD: 12.3). The study that reported a decline in guideline concordance was conducted in a secondary care hospital in the United Arab Emirates; this study assessed the impact of a 3-month intervention to develop and implement a new treatment guideline and conduct educational lectures for physicians for adopting the guidelines into their practice. The proportion of patients who received antimicrobial treatment that complied with guidelines regarding the timing of discontinuation fell from 29.7 to 8.3% (Odds ratio: 0.2, 95% CI: 0.1-0.4; Chi square p<0.001).

#### Theme 3: Antimicrobial stewardship

We extracted AMS data from 20 studies; 9 (45%) of which demonstrated significant improvements in AMS practices and 1 (5%) that reported a significant reduction in AMS practices (Fig 4). Two of three studies (66.7%) that assessed changes in intravenous to oral stepdown reported pre- to post-intervention improvements in conversion rates while 7 of 18 studies (38.8%) that assessed changes in the proportion of patients or cases where a culture or clinical diagnostic test was obtained reported significant pre- to post-increases. Of the studies the reported significant improvements in AMS, 3 were conducted in lower-middle-income countries (Egypt, India, Vietnam), 2 in upper-middle-income countries (China, South Africa), and 4 in high-income countries (Canada; USA, n=4). Most studies assessed bundled interventions (n=6, 66.7%) that were implemented in tertiary or academic health facilities (n=6, 66.7%). Four interventions involved the development and/or dissemination of treatment guidelines, protocols, or policies; 4 involved education or training sessions; 4 included the introduction of or changes to antibiotic order forms or clinical-support tools; 3 interventions involved audit or review and feedback; 1 study included the development of a facility antibiogram. Intervention durations ranged from 2 months to 24 months. One study assessed the impact of a one-year audit/review and feedback intervention on prescribers at a pediatric hospital in Iran obtaining blood cultures to aid in diagnosis and found that the proportion of patients whose care included a blood culture for diagnosis declined from 23.9% pre-intervention to 4.4% post-intervention (OR: 0.1, 95% CI: 0.04-0.5; Chi square p<0.001). The study also reported non-significant pre- to post-intervention declines in the proportion of patients whose diagnosis was aided by tracheal and urine cultures.

**Fig 4.**
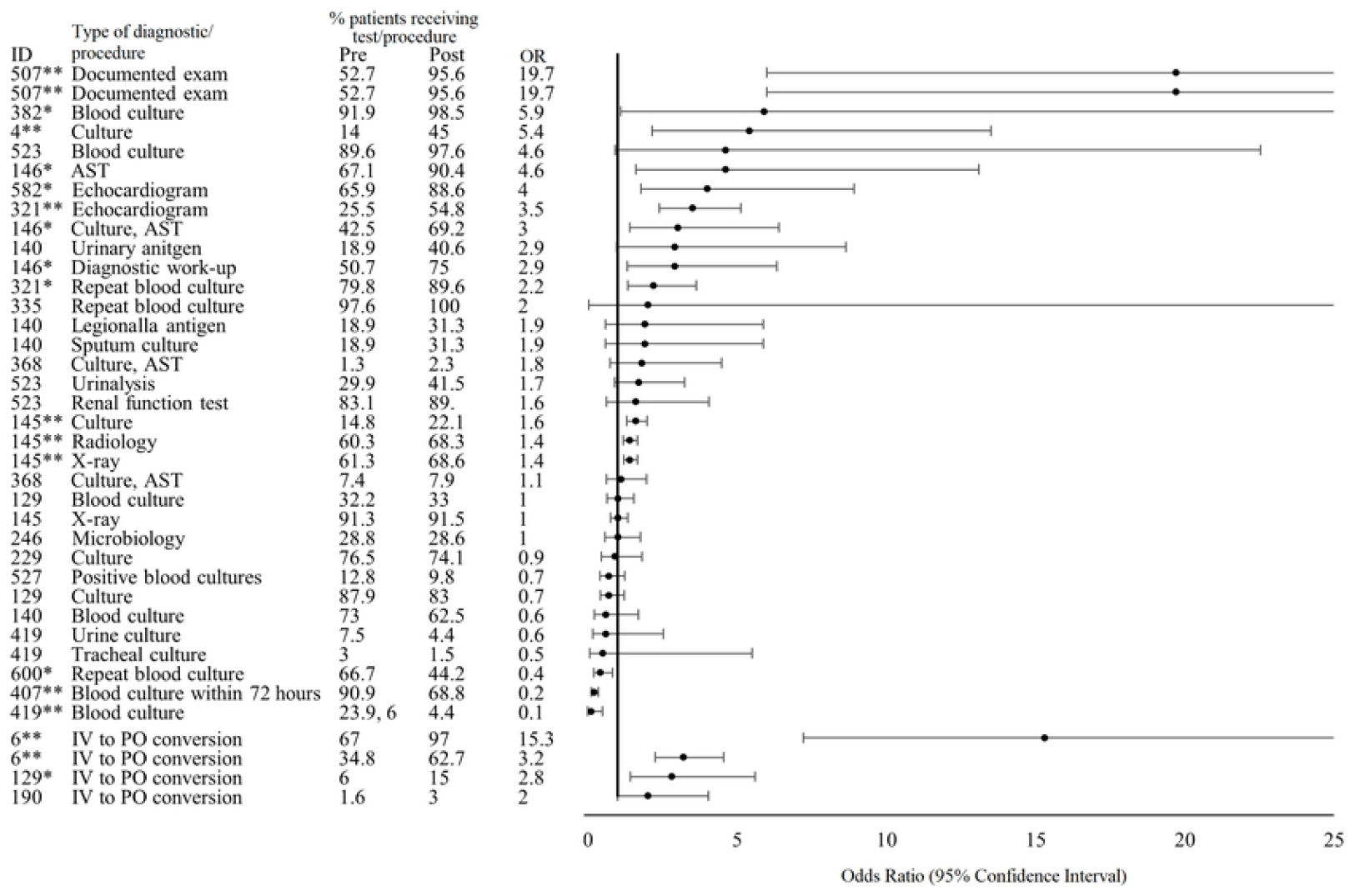
Effect of interventions on the utilization of clinical diagnostic and laboratory tests/cultures and intravenous to oral conversion rates. *Indicates significance <0.05. **Indicates significance <0.001. Abbreviations: AST-antimicrobial susceptibility testing, IV-intravenous, PO-per os (by mouth/oral).

#### Theme 4: Antimicrobial resistance

Of the studies that met our inclusion criteria and reported on our primary outcomes of interest, 11 reported on our secondary outcome of pre- to post-intervention changes in AMR (Fig 5). Across these studies, data was reported for 60 unique organism-antimicrobial combinations. Five studies (45.5%) reported significant decreases in the proportion of isolates that were resistant to antibiotics or the proportion of patients with drug-resistant infections across 17 antimicrobial-organism combinations while 3 studies (27.3%) reported both significant increases and decreases in resistance rates for different antimicrobial-organism combinations. All 8 studies reporting AMR data were conducted in high-income countries (Canada, Israel, France, Japan, Korea, Spain, and the USA; n=2). The 5 studies that reported only decreases in resistance rates were conducted in either acute care hospitals, nursing homes, public hospitals, or teaching hospitals (n=2); most of the studies tested single activity interventions (n=4, 80%) including introducing a treatment management protocol, and audit or review and feedback (n=3). The bundled intervention consisted of the implementation of an AMS toolkit and subsequent education and an antibiotic mobile team for daily AMS coordination.

**Fig 5.**
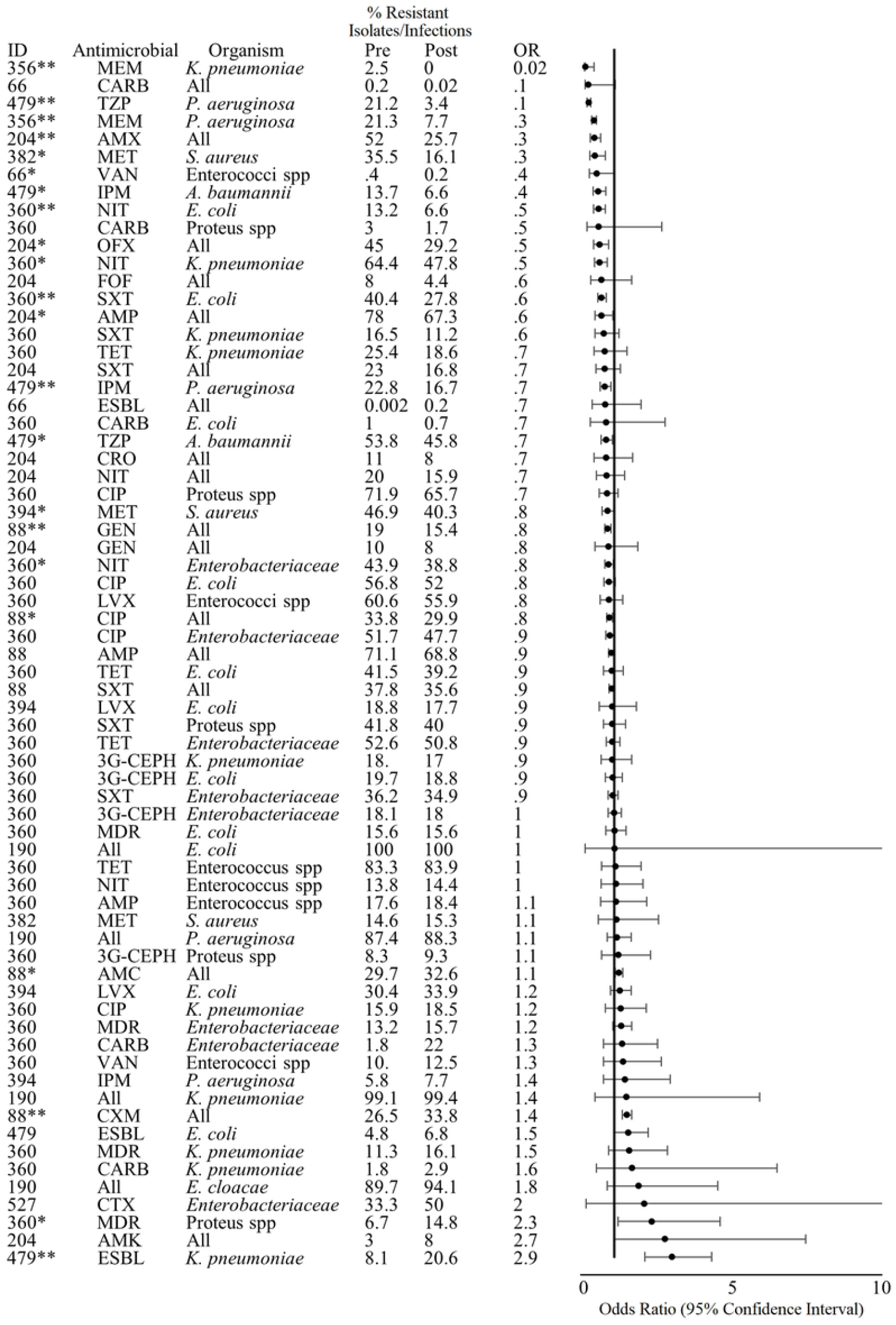
Effect of interventions on the proportion of isolates/infections resistant to various antimicrobials. *Indicates significance <0.05. **Indicates significance <0.001. Antimicrobial Abbreviations: AMK-amikacin, AMX-amoxicillin, AMC-amoxicillin-clavulanic acid, AMP-ampicillin, CARB-carbapenems, CTX-cefotaxime, CIP-ciprofloxacin, FOF-Fosfomycin, IPM-imipenem, LVX-levofloxacin, MDR-multi-drug resistant, MEM-meropenem, MET-methicillin, NIT-nitrofurantoin, TZP-piperacillin-tazobactam, SXT-trimethoprim-sulfamethoxazole, 3G-CEPH-third-generation cephalosporins.

The 3 studies that reported a combination of significant pre- to post-intervention changes were conducted in nursing homes or tertiary/teaching hospitals (n=2); two of these interventions consisted of bundled AMS activities. One intervention described by Ziv-On *et al*. consisted of review and feedback, education, and restricted use of certain antibiotics that required pre-prescription authorization (41). There were significant pre- to post-intervention decreases in the proportion of cultured isolates resistant to gentamycin (19% to 15.4%, OR: 0.8, 95% CI: 0.677-0.888, Chi-square p<0.001) and ciprofloxacin (33.8% to 29.9%, OR: 0.8, 95% CI: 0.7-0.9, Chi-square p=0.001) and significant increases in the proportion of isolates resistant to cefuroxime (26.5 to 33.8%, OR: 1.4, Chi-square p<0.001) and amoxicillin-clavulanic acid (29.7 to 32.6%, OR: 1.1, 95% CI: 1.03-1.3, Chi-square p=0.007). There were decreases in average quarterly use of aminoglycoside (gentamycin) from 2.3 (SD: 0.2) to 1.7 (SD: 0.3) daily defined doses/100 days of admission, quinolones (ciprofloxacin) 10.9 (SD: 0.7) to 6.71 (SD: 1.8), second-generation cephalosporins (cefuroxime) from 11.3 (SD: 0.7) to 8.6 (SD: 2.7), and amoxicillin-clavulanic acid from 19.8 (SD: 4.7) to 9.9 (SD: 3.7).

The second bundled study by Tandan *et al*. consisted of the introduction of a treatment guideline, education, and audit and feedback (42). There were significant decreases in the proportion of cultured *E. coli* isolates resistant to nitrofurantoin (13.2 to 6.6%, OR: 0.5, 95% CI: 0.3-0.7. Chi-square p<0.001) and trimethoprim-sulfamethoxazole (40.4 to 27.8%, OR: 0.6, 95% CI: 0.4-0.7, Chi-square p<0.001) and *Klebsiella pneumoniae* and Enterobacteriaceae to nitrofurantoin (64.4 to 47.8, OR: 0.5, 95% CI: 0.334-0.768; 43.9 to 38.8%, OR: 0.8, 95% CI: 0.7-1.0, respectively) but a significant increase in the proportion of *Proteus* spp. isolates that were multi-drug resistant (6.7 to 14.8%, OR: 2.3, 95% CI: 1.1-4.6, Chi-square p=0.02). The study did not report data on changes in AMU.

The third intervention described by Kim *et al*. tested the impact of restricting prescription of third-generation cephalosporin through a computerized antimicrobial prescription program (43). There were significant decreases in the proportion of *P. aeruginosa* isolates resistant to imipenem (22.8% to 16.7%, OR: 0.6, 95% CI: 0.5-0.9) and *A. baumannii* isolates resistant to imipenem (13.7% to 6.6%, OR: 0.4, 95% CI: 0.3-0.7) and trimethoprim-sulfamethoxazole (53.8% to 45.8%, OR: 0.7, 95% CI: 0.6-0.9) but a significant increase in the proportion of extended-spectrum beta-lactamase (ESBL)-producing *K. pneumoniae* isolates (8.1% to 20.6%, OR: 2.9, 95% CI: 2.0-4.3). There was no significant change in carbapenem (imipenem) or beta-lactamase inhibitor use, and data on sulfonamide (trimethoprim-sulfamethoxazole) use was not reported.

#### Theme 5: Clinical Outcomes

We extracted clinical outcome data from 57 of 301 total studies (Fig 6). Of three studies that reported the proportion of patients who were admitted to the intensive care unit before and after the intervention, only one reported a statistically significant change, a decrease from 44.5% to 35.5% of patients (OR: 0.7, 95% CI: 0.5-1.0) (44). Three separate studies reported the proportion of patients who experienced an adverse event before and after the intervention, and one reported a statistically significant change, an increase from 1.1% to 10.0% (OR: 9.9, 95% CI: 8.4-11.6) (45). Of 41 total studies that reported data on patient mortality before and after AMS interventions, 6 (14.6%) reported statistically significant decreases in the proportion of patients who died while 2 (4.8%) reported statistically significant increases in the proportion of patients who died; all other studies reported no significant change in mortality rates. Of 18 studies that reported on readmission rates, one study reported a significant increase and one a significant decrease in readmission after the intervention compared to before. Six of 8 studies (75%) that recorded the proportion of patients who experienced a recurrence or re-infection reported no significant pre- to post-intervention while two (25%) reported significant decreases. Finally, three studies reported on the proportion of patients experiencing treatment failure before and after interventions with only one reporting a significant pre- to post-intervention change, a decrease from 33.3% to 14.0% (OR: 0.3, 95% CI: 0.2-0.5) (46).

**Fig 6.**
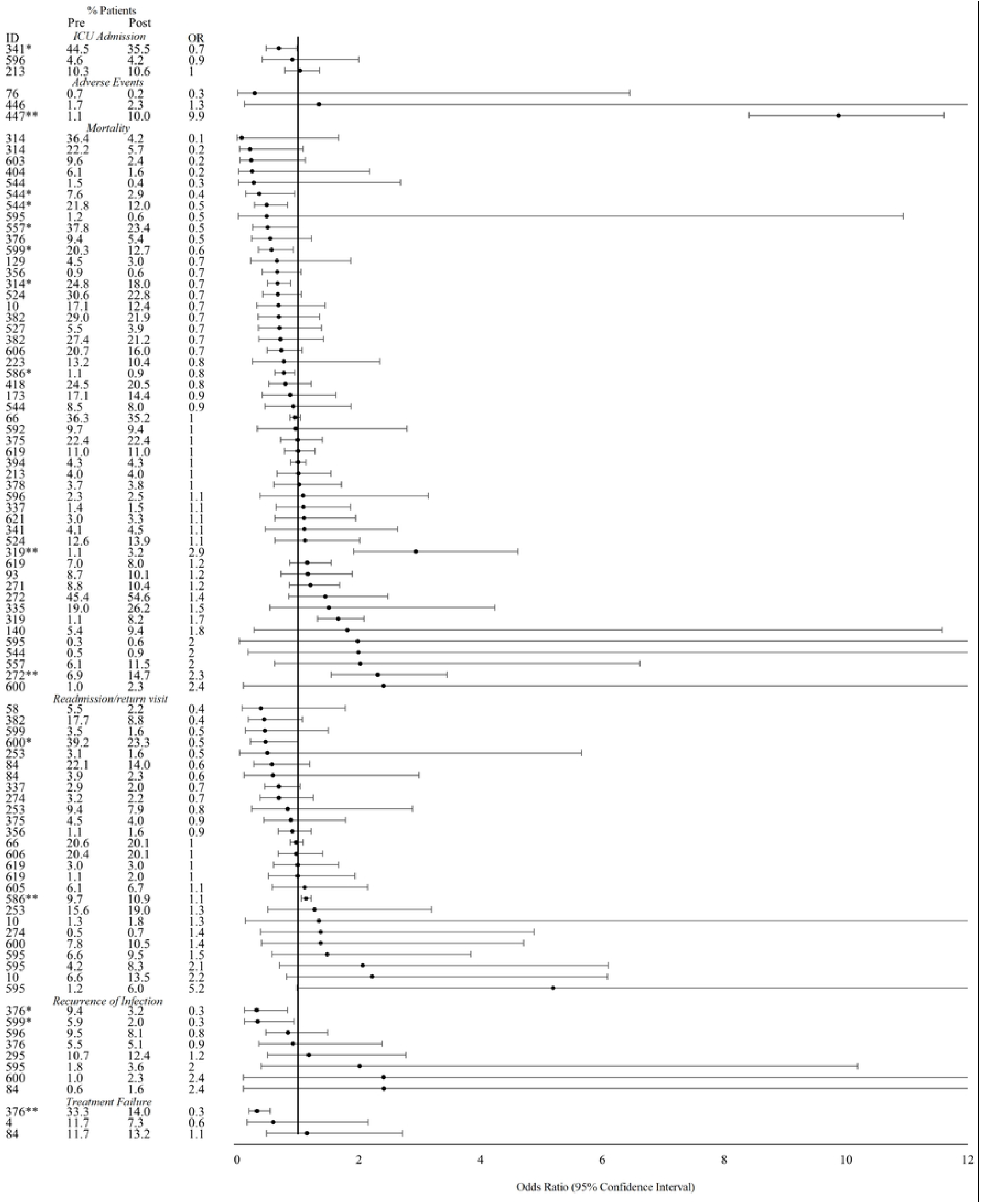
Effect of interventions on clinical outcomes (ICU admission rate, adverse events, mortality, readmission or return visit to a health facility, recurrence of infection, and treatment failure). *Indicates significance <0.05. **Indicates significance <0.001. Abbreviations: ICU-intensive care unit.

Based on univariable t-tests and multivariable linear regression, there were no statistically significant (p<0.05) associations between pre- to post-intervention absolute differences and intervention type, duration, or setting of intervention across any of our study outcomes in Themes 1-5.

## Discussion

Overall, there was a paucity of studies that assessed the impact of behavior-change interventions on AMU, AMS, and AMR in the animal health sector; further evidence is needed to understand best practices in these settings. A minority of studies conducted in the human health sector (35.7%) reported significant (p<0.05) pre- to post-intervention decreases in AMU while a majority (73.7%) reported significant improvements in adherence of antimicrobial therapies to local, national, or international guidelines. Of 20 studies, 45% demonstrated significant improvements in AMS practices while only 1 study reported a significant reduction in AMS practices. Eleven studies reported on pre- to post-intervention changes in the AMR burden, all of which took place in high-income countries. Five of those studies (45.5%) reported significant decreases in the proportion of isolates that were resistant to antibiotics or the proportion of patients with drug-resistant infections across 17 antimicrobial-organism combinations; 3 studies (27.3%) reported both significant increases and decreases in resistance rates for different antimicrobial-organism combinations. Generally, significant pre- to post-intervention changes in clinical outcomes – including ICU admission, mortality, adverse events, reinfection, mortality, and treatment failure rates – were infrequent with studies more commonly reporting improvements across those outcomes. Our study found no intervention characteristics that were associated with overall success the various thematic areas in any setting or subsetting included.

This study had several limitations. We limited our review to peer-reviewed studies which may have overlooked a significant number of ongoing interventions in various settings to improve AMR, AMU, and AMS and may have also introduced a publication bias where successful studies are more likely to be more published than unsuccessful interventions. Moreover, we only considered interventions that targeted behavior change among stakeholder groups involved in the use or prescription of antimicrobials thereby overlooking other important interventions such as those that aim to increase awareness and knowledge of AMR, AMU, and AMS; those that effect funding levels or policies; those that change access to clinically appropriate antimicrobials or other critical diagnostic and laboratory testing infrastructure; among other interventions and efforts that can effectively improve AMU and AMS and reduce AMR amongst human and animal populations. There were also limitations in the studies included in our final analysis. First, the majority of studies took place in developed countries; given the disparities in healthcare infrastructure, human resources, and AMR burden in high-versus low-resource settings, our findings may not be globally applicable. Notably, adherence to guidelines was the most improved intervention outcome in the human health sector; however, in many LMICs, local or national clinical treatment guidelines are often unavailable or outdated. A 2021 review of antimicrobial treatment guidelines in Africa found that only 20 of the 55 Africa Union member states had national guidelines at the time of the study, and only 10 member states had updated those guidelines since 2015 (47).

There were also limitations in the quality of evidence provided by the included studies. Few interventions utilized random allocation of treatment groups; most interventions were assessed using pre/post observational studies with no control group. The duration of interventions varied widely with many only assessing short-term impacts of interventions. Moreover, few studies described the theories underpinning behavior-change interventions which may improve effectiveness (48). Our quality assessment examined the risk of selection bias, study design, data collection methods, intervention integrity, and analyses, among other criteria. Overall, 4 of 11 studies (36.4%) that described interventions in the animal health or agriculture sectors and 129 of the 290 studies (44.5%) that described interventions in the human health sector were determined to have strong overall quality; therefore, there is room for improvement in overall study quality.

Overall, there is a need to increase the evidence base of behavior-change interventions regarding AMU, AMS, and AMR in the animal health and LMIC settings. Due to heterogeneity in the study settings and designs, it is difficult to identify which practices are most impactful across the board. There is a need to systematize approaches to assessing the impact of interventions and for studies to collect and report data representing the multiple ways AMR and AMS impacts clinical practice and outcomes; for example, studies should consider not only overall AMU rates but the clinical appropriateness of therapy; adherence to local, national, or international guidelines; clinical outcomes, AMS practices, cost and duration of patient stays; and clinical outcomes. There is a need to systematically assess interventions to allow for a thorough comparative analysis so that best practices in various settings can be identified for scaling these efforts.

## Data Availability

All data is provided in the supplementary materials.

## Acknowledgements

We thank Nathan Freeman for assistance with the quality assessment.

## References

1. World Health Organization. Antimicrobial resistance [Internet]. Geneva; 2021 Nov [cited 2022 Sep 19]. Available from: https://www.who.int/news-room/fact-sheets/detail/antimicrobial-resistance

2. World Bank Group. Drug-Resistant Infections-A Threat to Our Economic Future. Drug-Resistant Infections. Washington, DC; 2017.

3. George A. Antimicrobial resistance, trade, food safety and security. One Heal [Internet]. 2018 Jun 1 [cited 2022 Oct 1];5:6. Available from: /pmc/articles/PMC5725214/

4. Murray CJ, Ikuta KS, Sharara F, Swetschinski L, Robles Aguilar G, Gray A, et al. Global burden of bacterial antimicrobial resistance in 2019: a systematic analysis. Lancet [Internet]. 2022 Feb 12 [cited 2022 Sep 6];399(10325):629–55. Available from: http://www.thelancet.com/article/S0140673621027240/fulltext

5. Nelson RE, Hatfield KM, Wolford H, Samore MH, Scott RD, Reddy SC, et al. National Estimates of Healthcare Costs Associated With Multidrug-Resistant Bacterial Infections Among Hospitalized Patients in the United States. Clin Infect Dis [Internet]. 2021 Jan 29 [cited 2022 Sep 6];72(Supplement_1):S17–26. Available from: https://academic.oup.com/cid/article/72/Supplement_1/S17/6123350

6. Magnusson U, Moodley A, Osbjer K. Antimicrobial resistance at the livestock-human interface: implications for Veterinary Services. Rev Sci Tech Off Int Epiz. 2021;40(2).

7. Cooper B, Okello WO. An economic lens to understanding antimicrobial resistance: disruptive cases to livestock and wastewater management in Australia. Aust J Agric Resour Econ [Internet]. 2021 Oct 1 [cited 2022 Oct 1];65(4):900–17. Available from: https://onlinelibrary.wiley.com/doi/full/10.1111/1467-8489.12450

8. Klein EY, Van Boeckel TP, Martinez EM, Pant S, Gandra S, Levin SA, et al. Global increase and geographic convergence in antibiotic consumption between 2000 and 2015. Proc Natl Acad Sci U S A [Internet]. 2018 Apr 10 [cited 2022 Sep 19];115(15):E3463–70. Available from: https://www.pnas.org/doi/abs/10.1073/pnas.1717295115

9. Klein EY, Milkowska-Shibata M, Tseng KK, Sharland M, Gandra S, Pulcini C, et al. Assessment of WHO antibiotic consumption and access targets in 76 countries, 2000–15: an analysis of pharmaceutical sales data. Lancet Infect Dis [Internet]. 2021 Jan 1 [cited 2022 Oct 1];21(1):107–15. Available from: http://www.thelancet.com/article/S1473309920303327/fulltext

10. J G, M B, B W. Antibiotic prescribing practice and adherence to guidelines in primary care in the Cape Town Metro District, South Africa. S Afr Med J [Internet]. 2018 Mar 28 [cited 2022 Oct 1];108(4):304. Available from: https://pubmed.ncbi.nlm.nih.gov/29629681/

11. Wang J, Wang P, Wang X, Zheng Y, Xiao Y. Use and prescription of antibiotics in primary health care settings in China. JAMA Intern Med [Internet]. 2014 Dec 1 [cited 2022 Oct 1];174(12):1914–20. Available from: https://pubmed.ncbi.nlm.nih.gov/25285394/

12. Sarwar MR, Saqib A, Iftikhar S, Sadiq T. Antimicrobial use by WHO methodology at primary health care centers: A cross sectional study in Punjab, Pakistan 11 Medical and Health Sciences 1117 Public Health and Health Services. BMC Infect Dis [Internet]. 2018 Sep 29 [cited 2022 Oct 1];18(1):1–9. Available from: https://bmcinfectdis.biomedcentral.com/articles/10.1186/s12879-018-3407-z

13. Versporten A, Bielicki J, Drapier N, Sharland M, Goossens H, Calle GM, et al. The worldwide antibiotic resistance and prescribing in european children (ARPEC) point prevalence survey: Developing hospital-quality indicators of antibiotic prescribing for children. J Antimicrob Chemother. 2016;71(4):1106–17.

14. Tiseo K, Huber L, Gilbert M, Robinson TP, Van Boeckel TP. Global Trends in Antimicrobial Use in Food Animals from 2017 to 2030. Antibiotics [Internet]. 2020 Dec 1 [cited 2022 Sep 19];9(12):1–14. Available from: /pmc/articles/PMC7766021/

15. Schar D, Klein EY, Laxminarayan R, Gilbert M, Van Boeckel TP. Global trends in antimicrobial use in aquaculture. Sci Reports 2020 101 [Internet]. 2020 Dec 14 [cited 2022 Oct 1];10(1):1–9. Available from: https://www.nature.com/articles/s41598-020-78849-3

16. Frost I, Craig J, Joshi J, Faure K, Laxminarayan R. Access Barriers to Antibiotics. Washington, DC; 2019.

17. Owens RC. Antimicrobial stewardship: concepts and strategies in the 21st century. Diagn Microbiol Infect Dis [Internet]. 2008 May [cited 2022 Oct 1];61(1):110–28. Available from: https://pubmed.ncbi.nlm.nih.gov/18384997/

18. MacDougall C, Polk RE. Antimicrobial stewardship programs in health care systems. Clin Microbiol Rev [Internet]. 2005 Oct [cited 2022 Oct 1];18(4):638–56. Available from: https://pubmed.ncbi.nlm.nih.gov/16223951/

19. Garau J, Bassetti M. Role of pharmacists in antimicrobial stewardship programmes. Int J Clin Pharm [Internet]. 2018 Oct 1 [cited 2022 Oct 1];40(5):948–52. Available from: https://pubmed.ncbi.nlm.nih.gov/30242589/

20. Moher D, Liberati A, Tetzlaff J, Altman DG. Preferred reporting items for systematic reviews and meta-analyses: the PRISMA statement. BMJ [Internet]. 2009 Jul 21 [cited 2022 Oct 10];339(7716):332–6. Available from: https://www.bmj.com/content/339/bmj.b2535

21. National Library of Medicine. Pubmed.gov [Internet]. 1996. Available from: https://pubmed.ncbi.nlm.nih.gov/

22. Clarivate. Web of Science. 1997; Available from: https://access.clarivate.com/login?app=wos&alternative=true&shibShireURL=https:%2F%2Fwww.webofknowledge.com%2F%3Fauth%3DShibboleth&shibReturnURL=https:%2F%2Fwww.webofknowledge.com%12F&roaming=truehttps://access.clarivate.com/login?app=wos&alternative=true&shibShireURL=https://%2F%2Fwww.webofknowledge.com%2F%3Fauth%3DShibboleth&shibReturnURL=https://%2F%2Fwww.web

23. Elsevier. Embase. 1947; Available from: https://www.embase.com/

24. Centre for Agriculture and Bioscience International. 1910; Available from: https://www.cabi.org/

25. Cochrane Library. Cochrane Database of Systematic Reviews. Available from: https://www.cochranelibrary.com/cdsr/about-cdsr

26. The World Bank. World Bank Country and Lending Groups [Internet]. 2022 [cited 2022 Oct 1]. Available from: https://datahelpdesk.worldbank.org/knowledgebase/articles/906519-world-bank-country-and-lending-groups

27. Effective Public Health Practice Project. Quality Assessment Tool for Quantitative Studies. 1998;

28. Phu DH, Cuong N Van, Truong DB, Kiet BT, Hien VB, Thu HTV, et al. Reducing Antimicrobial Usage in Small-Scale Chicken Farms in Vietnam: A 3-Year Intervention Study. Front Vet Sci. 2021 Jan 28;7:1244.

29. Hubbuch A, Peter R, Willi B, Hartnack S, Müntener C, Naegeli H, et al. Comparison of antimicrobial prescription patterns in calves in Switzerland before and after the launch of online guidelines for prudent antimicrobial use. BMC Vet Res [Internet]. 2021 Dec 1 [cited 2022 Oct 1];17(1):1–14. Available from: https://bmcvetres.biomedcentral.com/articles/10.1186/s12917-020-02704-w

30. Gerber M, Dürr S, Bodmer M. Reducing Antimicrobial Use by Implementing Evidence-Based, Management-Related Prevention Strategies in Dairy Cows in Switzerland. Front Vet Sci [Internet]. 2021 Jan 18 [cited 2022 Oct 1];7. Available from: https://pubmed.ncbi.nlm.nih.gov/33537355/

31. Verliat F, Hemonic A, Chouet S, Le Coz P, Liber M, Jouy E, et al. An efficient cephalosporin stewardship programme in French swine production. Vet Med Sci [Internet]. 2021 Mar 1 [cited 2022 Oct 1];7(2):432–9. Available from: https://onlinelibrary.wiley.com/doi/full/10.1002/vms3.377

32. Collineau L, Rojo-Gimeno C, Léger A, Backhans A, Loesken S, Nielsen EO, et al. Herd-specific interventions to reduce antimicrobial usage in pig production without jeopardising technical and economic performance. Prev Vet Med. 2017 Sep 1;144:167–78.

33. Speksnijder DC, Graveland H, Eijck IAJM, Schepers RWM, Heederik DJJ, Verheij TJM, et al. Effect of structural animal health planning on antimicrobial use and animal health variables in conventional dairy farming in the Netherlands. J Dairy Sci [Internet]. 2017 Jun 1 [cited 2022 Oct 1];100(6):4903–13. Available from: https://pubmed.ncbi.nlm.nih.gov/28390724/

34. Jensen VF, de Knegt L V., Andersen VD, Wingstrand A. Temporal relationship between decrease in antimicrobial prescription for Danish pigs and the “Yellow Card” legal intervention directed at reduction of antimicrobial use. Prev Vet Med [Internet]. 2014 Dec 1 [cited 2022 Oct 1];117(3–4):554–64. Available from: https://pubmed.ncbi.nlm.nih.gov/25263135/

35. Singleton DA, Rayner A, Brant B, Smyth S, Noble PJM, Radford AD, et al. A randomised controlled trial to reduce highest priority critically important antimicrobial prescription in companion animals. Nat Commun 2021 121 [Internet]. 2021 Mar 11 [cited 2022 Oct 1];12(1):1–14. Available from: https://www.nature.com/articles/s41467-021-21864-3

36. Lehner C, Hubbuch A, Schmitt K, Schuepbach-Regula G, Willi B, Mevissen M, et al. Effect of antimicrobial stewardship on antimicrobial prescriptions for selected diseases of dogs in Switzerland. J Vet Intern Med [Internet]. 2020 Nov 1 [cited 2022 Oct 1];34(6):2418–31. Available from: https://pubmed.ncbi.nlm.nih.gov/33112451/

37. Hubbuch A, Schmitt K, Lehner C, Hartnack S, Schuller S, Schüpbach-Regula G, et al. Antimicrobial prescriptions in cats in Switzerland before and after the introduction of an online antimicrobial stewardship tool. BMC Vet Res [Internet]. 2020 Jul 3 [cited 2022 Oct 1];16(1):1–13. Available from: https://bmcvetres.biomedcentral.com/articles/10.1186/s12917-020-02447-8

38. Hopman NEM, Portengen L, Hulscher MEJL, Heederik DJJ, Verheij TJM, Wagenaar JA, et al. Implementation and evaluation of an antimicrobial stewardship programme in companion animal clinics: A stepped-wedge design intervention study. PLoS One [Internet]. 2019 Nov 1 [cited 2022 Oct 1];14(11). Available from: https://pubmed.ncbi.nlm.nih.gov/31738811/

39. Leis JA, Palmay L, Ho G, Raybardhan S, Gill S, Kan T, et al. Point-of-Care β-Lactam Allergy Skin Testing by Antimicrobial Stewardship Programs: A Pragmatic Multicenter Prospective Evaluation. Clin Infect Dis [Internet]. 2017 Oct 1 [cited 2022 Nov 27];65(7):1059–65. Available from: https://pubmed.ncbi.nlm.nih.gov/28575226/

40. Yogo N, Shihadeh K, Young H, Calcaterra SL, Knepper BC, Burman WJ, et al. Intervention to Reduce Broad-Spectrum Antibiotics and Treatment Durations Prescribed at the Time of Hospital Discharge: A Novel Stewardship Approach. Infect Control Hosp Epidemiol [Internet]. 2017 May 1 [cited 2022 Nov 27];38(5):534–41. Available from: https://pubmed.ncbi.nlm.nih.gov/28260538/

41. Ziv-On E, Friger MD, Saidel-Odes L, Borer A, Shimoni O, Nikonov A, et al. Impact of an Antibiotic Stewardship Program on the Incidence of Resistant Escherichia coli: A Quasi-Experimental Study. Antibiot (Basel, Switzerland) [Internet]. 2021 Feb 1 [cited 2022 Nov 27];10(2):1–10. Available from: https://pubmed.ncbi.nlm.nih.gov/33578840/

42. Tandan M, Sloane PD, Ward K, Weber DJ, Vellinga A, Kistler CE, et al. Antimicrobial resistance patterns of urine culture specimens from 27 nursing homes: Impact of a two-year antimicrobial stewardship intervention. Infect Control Hosp Epidemiol [Internet]. 2019 Jul 1 [cited 2022 Nov 27];40(7):780–6. Available from: https://pubmed.ncbi.nlm.nih.gov/31057141/

43. Kim JY, Sohn JW, Park DW, Yoon YK, Kim YM, Kim MJ. Control of extended-spectrum {beta}-lactamase-producing Klebsiella pneumoniae using a computer-assisted management program to restrict third-generation cephalosporin use. J Antimicrob Chemother [Internet]. 2008 Aug [cited 2022 Nov 27];62(2):416–21. Available from: https://pubmed.ncbi.nlm.nih.gov/18413317/

44. Shemanski S, Bennett N, Essmyer C, Kennedy K, Buchanan DM, Warnes A, et al. Centralized Communication of Blood Culture Results Leveraging Antimicrobial Stewardship and Rapid Diagnostics. Open Forum Infect Dis [Internet]. 2019 Sep 1 [cited 2022 Nov 27];6(9). Available from: https://academic.oup.com/ofid/article/6/9/ofz321/5532664

45. Shawki MA, AlSetohy WM, Ali KA, Ibrahim MR, El-Husseiny N, Sabry NA. Antimicrobial stewardship solutions with a smart innovative tool. J Am Pharm Assoc (2003) [Internet]. 2021 Sep 1 [cited 2022 Nov 27];61(5):581-588.e1. Available from: https://pubmed.ncbi.nlm.nih.gov/33962893/

46. Niwa T, Yonetamari J, Hayama N, Fujibayashi A, Ito-Takeichi S, Suzuki K, et al. Clinical impact of matrix-assisted laser desorption ionization-time of flight mass spectrometry combined with antimicrobial stewardship interventions in patients with bloodstream infections in a Japanese tertiary hospital. Int J Clin Pract [Internet]. 2019 May 1 [cited 2022 Nov 27];73(5). Available from: https://pubmed.ncbi.nlm.nih.gov/30810264/

47. Craig J, Hiban K, Frost I, Kapoor G, Alimi Y, Varma JK. Comparison of national antimicrobial treatment guidelines, African Union. Bull World Health Organ [Internet]. 2022 Jan 1 [cited 2022 Apr 7];100(1):50. Available from: /pmc/articles/PMC8722630/

48. Korda H, Itani Z. Harnessing social media for health promotion and behavior change. Health Promot Pract [Internet]. 2013 Jan [cited 2022 Nov 29];14(1):15–23. Available from: https://pubmed.ncbi.nlm.nih.gov/21558472/

